# Metrics to relate COVID-19 wastewater data to clinical testing dynamics

**DOI:** 10.1101/2021.06.10.21258580

**Authors:** Amy Xiao, Fuqing Wu, Mary Bushman, Jianbo Zhang, Maxim Imakaev, Peter R Chai, Claire Duvallet, Noriko Endo, Timothy B Erickson, Federica Armas, Brian Arnold, Hongjie Chen, Franciscus Chandra, Newsha Ghaeli, Xiaoqiong Gu, William P Hanage, Wei Lin Lee, Mariana Matus, Kyle A McElroy, Katya Moniz, Steven F Rhode, Janelle Thompson, Eric J Alm

## Abstract

Wastewater surveillance has emerged as a useful tool in the public health response to the COVID-19 pandemic. While wastewater surveillance has been applied at various scales to monitor population-level COVID-19 dynamics, there is a need for quantitative metrics to interpret wastewater data in the context of public health trends. We collected 24-hour composite wastewater samples from March 2020 through May 2021 from a Massachusetts wastewater treatment plant and measured SARS-CoV-2 RNA concentrations using RT-qPCR. We show that the relationship between wastewater viral titers and COVID-19 clinical cases and deaths varies over time. We demonstrate the utility of three new metrics to monitor changes in COVID-19 epidemiology: (1) the ratio between wastewater viral titers and clinical cases (WC ratio), (2) the time lag between wastewater and clinical reporting, and (3) a transfer function between the wastewater and clinical case curves. We find that the WC ratio increases after key events, providing insight into the balance between disease spread and public health response. We also find that wastewater data preceded clinically reported cases in the first wave of the pandemic but did not serve as a leading indicator in the second wave, likely due to increased testing capacity. These three metrics could complement a framework for integrating wastewater surveillance into the public health response to the COVID-19 pandemic and future pandemics.

## Introduction

The coronavirus disease 2019 (COVID-19) pandemic, caused by severe acute respiratory syndrome coronavirus 2 (SARS-CoV-2), continues to affect all aspects of global life since its emergence in late 2019. Efforts to contain its spread have relied on public health measures like social distancing, stay-at-home orders, mandatory wearing of face coverings in public, and community-based SARS-CoV-2 testing (Nussbaumer-Streit et al., 2020). In the face of this public health emergency, multiple methods have been designed to assess the impact of SARS-CoV-2 on critical health infrastructure, implement rapid and robust individual testing, and predict potential surges to inform hospital preparedness and health policy makers.

Clinical surveillance data is the gold standard for evaluating the state of the pandemic. However, clinical data can be limited by clinical testing capacity and availability, human behavior, and the presence of asymptomatic infections. In contrast, wastewater surveillance, also known as wastewater-based epidemiology (WBE), is a potentially powerful strategy for near real-time monitoring of viral burden in the population, as it captures viral shedding from infected individuals, irrespective of clinical presentation. Such surveillance has been shown to detect SARS-CoV-2 in wastewater before widespread clinical reporting (Bar-Or et al., 2020; Kocamemi et al., 2020; Medema et al., 2020; Randazzo et al., 2020; Wurtzer et al., 2020). We and others have previously demonstrated that viral titers in wastewater were higher than expected from confirmed clinical cases (Wu et al., 2020b) and that they preceded clinical reporting of new cases and hospital admissions (Peccia et al., 2020; Randazzo et al., 2020; Wu et al., 2020a), suggesting that wastewater surveillance could be used as an early warning system. Wastewater surveillance has also been used in combination with clinical data to infer average population-level viral shedding dynamics (Schmitz et al., 2021; Wu et al., 2020a). Wastewater surveillance has been mostly implemented at municipal wastewater treatment plants, but as colleges and universities reopened, it has also been applied in dormitories as an early warning system to prevent large-scale outbreaks of COVID-19 (Betancourt et al., 2020; Gibas et al., 2021; Harris-Lovett et al., 2021; P. Liu et al., 2020).

Despite the diverse applications of wastewater surveillance as a detection and sentinel tool, there remains a need for standardized methods to quantitatively interpret wastewater data in the context of clinical trends and public health interventions. Such methods would provide a framework for incorporating wastewater-based metrics into the public health decision making toolkit, beyond solely observing the qualitative trends in viral titers. Here, we developed three new metrics and applied them to a 14-month-long time series of SARS-CoV-2 wastewater titers in Massachusetts. We show that these three metrics – (1) the ratio between wastewater viral titers and clinical cases (WC ratio), (2) the time lag between wastewater and clinical reporting, and (3) a transfer function between the wastewater and clinical case curves – aptly describe dynamic changes in the relationship between wastewater viral titers data and clinical data as the pandemic evolves and management strategies adapt. Our results show that wastewater data serves as an early warning for the first wave of the pandemic in Massachusetts but not the second, likely due to the large-scale ramp up of clinical testing availability.

## Results

### Wastewater viral titers mirror disease incidence and can depict the impact of social activities more sensitively than clinical cases

Analysis of our 14-month-long wastewater surveillance time series for SARS-CoV-2 viral titers spanning March 4, 2020 to May 13, 2021 showed two distinct waves of SARS-CoV-2 in the Boston Area. Similar trends appeared in wastewater viral levels and clinical cases: exponential rise from March to mid-April, a decline through July, a slow increase over the summer, followed by a sharper increase in the fall and second peak in the winter (Figure 1), indicating that wastewater viral titers generally mirrored trends in disease incidence.

**Figure 1.**
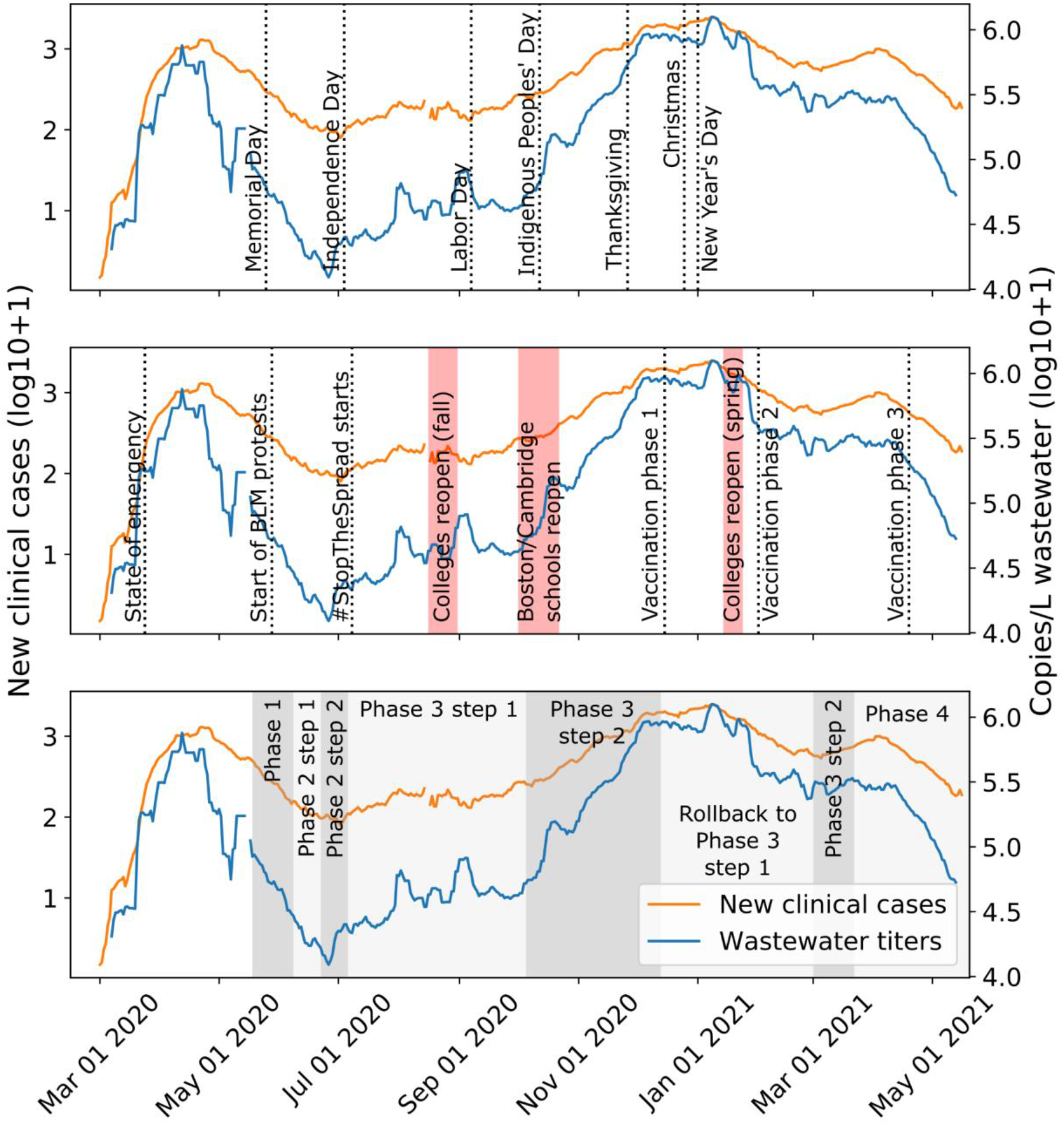
SARS-CoV-2 RNA titers in Massachusetts wastewater and new clinical cases. Seven-day averages of wastewater viral titers (blue) and new clinical cases reported for the three counties in the catchment (orange) (41-43). We marked major holidays (top), major social events (middle), and state reopening phases (bottom) in the three panels, respectively (Baker, 2021a, p. 2, 2021b, 2020a, 2020b, 2020c, 2020d, 2020e, 2020f, n.d.).

We compared wastewater and clinical data with dates of known policy changes and social gatherings in the Boston Area and noted key trends. For example, wastewater viral titers and clinical cases continued to decline overall after Memorial Day (May 25, 2020), despite the potential for large gatherings to celebrate the holiday (Figure 1). Similarly, the social justice protests during late May and June did not immediately spark an increase in clinical cases or wastewater viral titers (Figure 1). The start of Phase 2 Step 2 (Table S1) marked the start of a steady increase in wastewater viral titers and clinical cases during the summer (Figure 1). We also observed a steeper increase in wastewater viral titers and clinical cases after the Indigenous Peoples’ Day holiday (October 12, 2020). This increase continued through reopening Phase 3 Step 2 and peaked in late November to January around the time of Thanksgiving, Christmas, and New Year holidays, perhaps due to increased indoor gatherings (Figure 1).

However, trends in wastewater data differed from clinical data after some key events, suggesting a decoupling of wastewater and clinical trends which we hypothesize provides insight into dynamics of COVID-19 in the community. We observed a short peak in wastewater viral titers at the start of August, which was only slightly reflected in the clinical data (Figure 1). Similarly, after colleges and universities welcomed students back in late August/early September, we observed another peak in wastewater viral titers, but not in clinical cases (Figure 1). After the start of Phase 3 Step 2 reopening, wastewater viral titers increased steeply, while clinical cases had a shallower slope (Figure 1). The observation of distinct trends in wastewater titers over clinical data suggests that wastewater viral titers could indicate the impact of social activities more sensitively than clinical cases. This observation prompted us to derive a metric for detecting discordance between wastewater and clinical trends.

### Ratio between wastewater viral titer and clinical cases (WC ratio) may indicate changes in testing capacity or demographics of clinical cases

Wastewater surveillance captures all individuals who are shedding the virus regardless of disease manifestation, their access to testing, or representation in clinical case data. As such, wastewater trends are commonly used for benchmarking against clinical cases. However, there is a lack of a quantitative measure for comparing wastewater and clinical trends. Here, we propose using the ratio of wastewater viral titers to clinical cases (WC ratio) as a metric for detecting differences between wastewater and clinical trends.

Changes in the WC ratio can serve as an indicator of potential under- or over-estimation of disease incidence. Under-counting of clinical cases could occur when clinical tests are limited, when people are not seeking testing or cannot access convenient testing locations, or when the proportion of asymptomatic infections is high. Over-estimation of disease incidence could occur when rapid expansion of testing infrastructure allows many people who were infected in the prior weeks to get tested, so their results show up as new cases even though they were actually infected weeks ago. This situation could occur especially because throat and nasal swab PCR tests for SARS-CoV-2 can remain positive for up to 20 days after symptom onset (Wölfel et al., 2020).

Because viral titers will vary based on the number of people in the catchment, the magnitude and range of the WC ratio must be established for each catchment considered. When this ratio is high, it implies that the existing testing capacity has not kept pace with exponentially rising new cases, which nevertheless are detected in wastewater surveillance. A high WC ratio can occur in situations where wastewater viral titers are increasing but clinical cases are not rising equivalently, or conversely when wastewater viral titers are stable but clinical testing is decreasing. Conversely, a low WC ratio indicates that clinical tests are capturing the majority of infections reflected in wastewater viral titers. When this ratio is stable and low, it implies that the existing testing capacity is sufficient to assess the extent of new infections. Importantly, changes in the WC ratio relative to a stable baseline may provide early indications of changing epidemic dynamics. Positive changes may highlight new bursts of infections before they are captured in clinical data or identify periods where clinical tests are not capturing the full extent of new infections, while negative changes may highlight periods where clinical testing is over-estimating disease incidence when counting previously infected cases as new cases. Notably, this ratio has changed by approximately two orders of magnitude over the course of the pandemic, demonstrating its utility as a metric to assess the public health response.

At the beginning of the pandemic (March 2020), the ratio between wastewater viral titers (viral genome copies (GC) per L) and reported clinical cases was very high (>10^3 for the Boston Area), indicating that cases were likely undercounted due to extremely limited testing (Figure 2A). In fact, during March 2020, the seven-day average of new molecular tests administered per day in the state did not exceed 5,000 tests per day (Figure 2B). As testing ramped up throughout April and May, the WC ratio dipped by approximately two orders of magnitude (Figure 2A). In this phase, delayed clinical test results may have been “catching up” to the more instantaneous wastewater viral titers, which infected individuals had contributed to many days prior to getting their test results. Thus, these individuals were no longer contributing to wastewater viral titers (numerator of the WC ratio) but were now being counted as reported cases (denominator of the WC ratio), leading to a much lower ratio.

**Figure 2.**
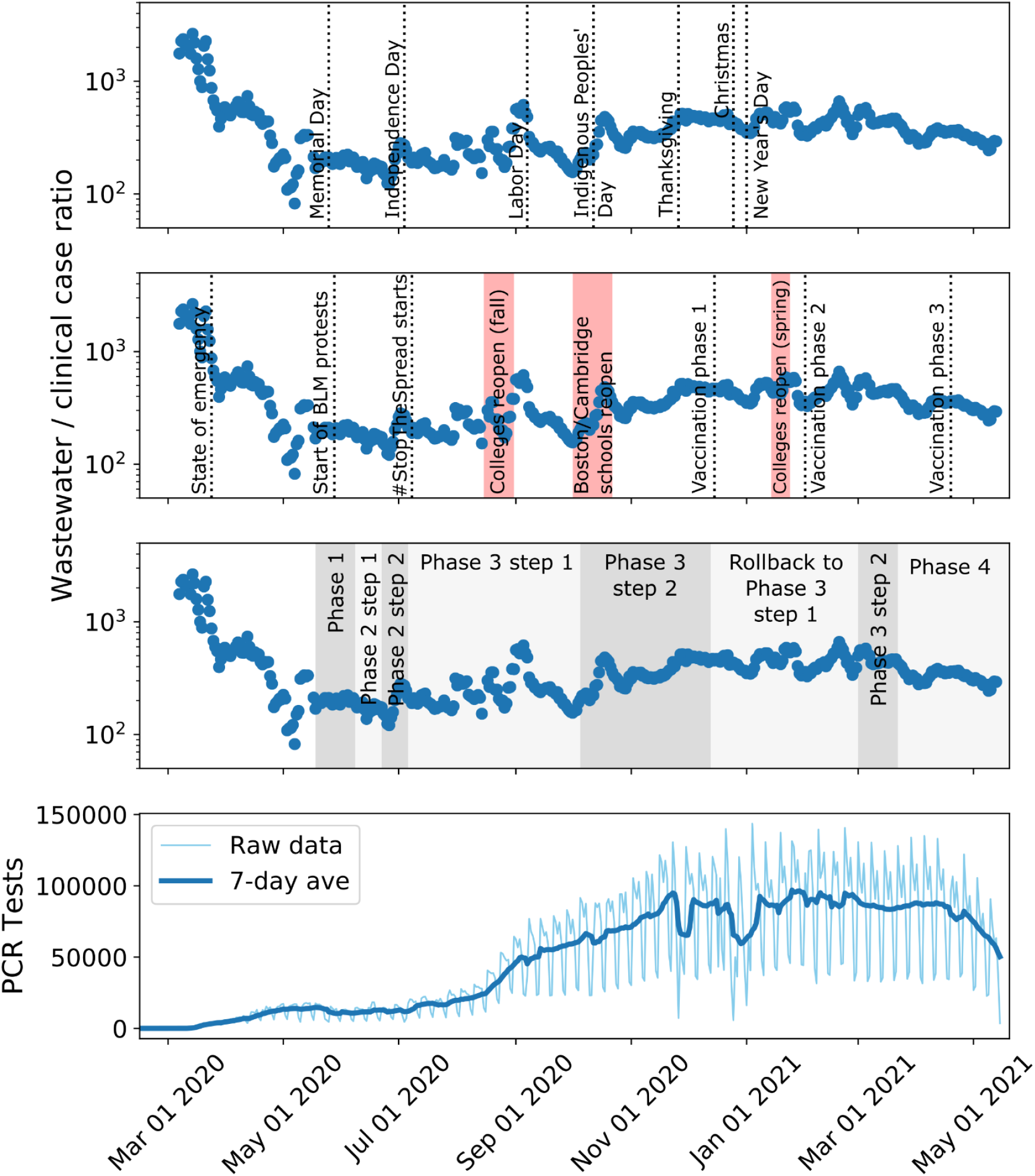
Ratio between wastewater viral titers and clinically reported new cases changes with testing availability. (A) Ratio between seven-day averages of wastewater viral concentration (genome copies/L) and clinically reported new cases changes over the course of the pandemic, with some spikes after key holidays, important events, and reopening phases. (B) PCR tests conducted each day in Massachusetts throughout the pandemic (Massachusetts Department of Public Health, 2021b).

As the public health response in Massachusetts began to ramp up over the summer and individual clinical testing became more available, the ratio between wastewater and clinical cases stayed fairly consistent between July 2020 and November 2020 (Figure 2A). During this time, Massachusetts’ clinical testing capacity became fully established, and percent positivity remained stable below 2% (Massachusetts Department of Public Health, 2021a). Such periods could be used to determine baseline WC ratios that indicate sufficient public health capacity.

Interestingly, even during the pronounced peak of the second wave (November 2020 - March 2021), the WC ratio did not spike, indicating that testing capacity was sufficient to capture the scope of exponentially rising new infections.

However, we did observe a few increases in the ratio even with fully established testing capacity, notably in early September and early October (Figure 2A). These increases in the WC ratio could be related to community events, such as reopening of businesses and universities, or to changes in testing availability. These increases could also be due to a combination of factors including shifts in population demographics. For example, college students returning for classes in late August/early September may have been more likely to be asymptomatic due to their younger age, and thus less likely to be reflected in clinical case counts (Leidman et al., 2021; Leidner et al., 2021). Similarly, after Phase 3 Step 2 reopening allowed bars and entertainment venues to open (Baker, 2021a), young adults may have had higher degrees of social contacts. In these instances, wastewater surveillance may have detected a silent and short-lived peak in community transmission that was missed by clinical surveillance. Importantly, these short increases became more apparent when analyzing the ratio between the two datasets.

### Wastewater lead time is affected by increased clinical testing

While the WC ratio could identify whether trends in clinical cases are concordant with trends in wastewater titers, it does not provide information on the timeliness of clinical reporting, which would be an important measure of the public health response. To address this gap, we introduce two methods to characterize this time lag between wastewater and clinical trends: (1) a model of the distribution of time lags for each new case, and (2) a transfer function to describe the relationship between the wastewater and clinical curves.

In our first metric to assess the time-varying relationship between wastewater data and clinical cases (Figure 3A), we used approximate Bayesian computation (ABC) to model the distribution of the time lag between when an infected person’s viral shedding is detectable in wastewater and when they receive a clinical test (Methods). In this model, we assume that the viral shedding detected in wastewater occurs on a single day. The assumption that wastewater reflects viral shedding early in infection is supported by animal models, meta-analysis of clinical data, and wastewater surveillance in dormitories (Bao et al., 2020; Hoffmann and Alsing, 2021; Schmitz et al., 2021). The date of clinical cases in this analysis corresponds to the date of specimen collection. Negative time lags indicate that wastewater signal precedes clinical testing and vice versa. To compare the relationship between wastewater data and clinical data during the first and second waves of the pandemic, we split the time series on August 15, the approximate midpoint between the end of the first wave and the start of the second wave.

**Figure 3.**
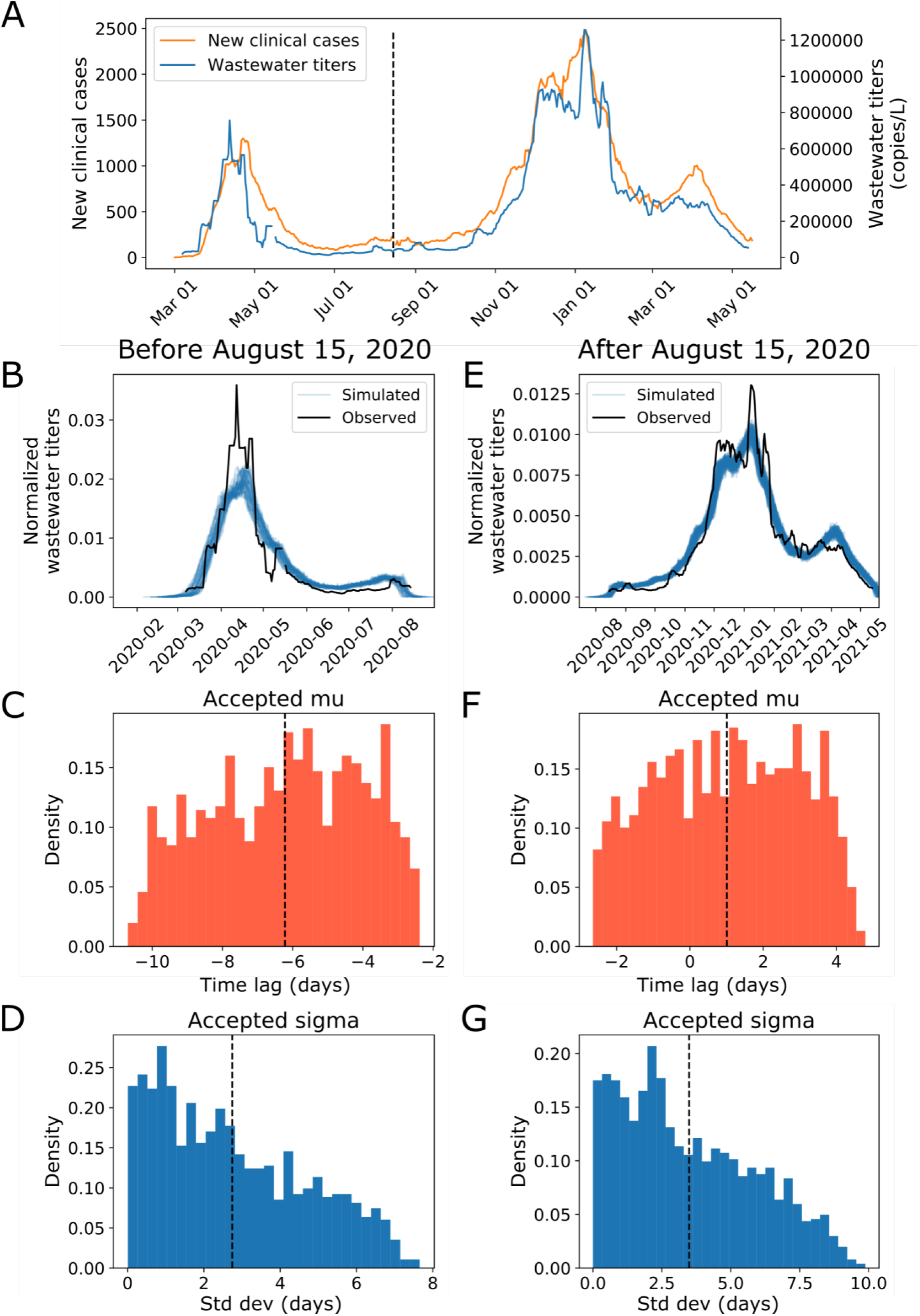
Modeling reveals that time delay between clinical case reporting and wastewater data changes over the course of the pandemic. We use Approximate Bayesian Computation to determine the distribution of the time lag between when a case shows up in the wastewater surveillance data and when they are clinically reported. (A) Seven-day averages of wastewater data and new clinical case data from Mass.gov shown on a linear scale (Massachusetts Department of Public Health, 2021b, 2020a, 2020b). (B, C, D) Modeling results on data before August 15, 2020. (B) Simulated vs observed wastewater viral titers for data before 8/15. (C) Accepted values for the mean time lag (−6.2 days, 95% CI: -10.1, -2.7) and (D) standard deviation (2.7 days, 95% CI: 0.1, 6.7) of the time lag for data before 8/15. We used 10,000 iterations with a distance threshold of 1.3e-5 and 11.1% of parameter sets were accepted. (E, F, G) Modeling results on data from August 15, 2020 and after. (E) Simulated vs observed wastewater viral titers for data after 8/15. (F) Accepted values for the mean time lag (1.0 days, 95% CI: -2.4, 4.2) and (G) standard deviation (3.5 days, 95% CI: 0.2, 8.4) of the time lag for data after 8/15. We used 10,000 iterations with a distance threshold of 9.5e-7 and 15.3% of parameter sets were accepted. Negative time lags indicate that wastewater signal precedes clinical case reporting and vice versa.

Before August 15, 2020, the wastewater signal preceded clinical cases by approximately 6.2 days (95% CI: -10.1, -2.7) with a standard deviation of 2.7 days (95% CI: 0.1, 6.7) (Figure 3B-D). This finding is consistent with our previous report that wastewater data preceded clinical data by 4-10 days (Wu et al., 2020a). After August 15, 2020, the wastewater signal was more in phase with clinical cases (mean time lag 1.0 days, 95% CI: -2.4, 4.2; standard deviation 3.5 days, 95% CI: 0.2, 8.4) (Figure 3E-G). We repeated the modeling while varying the date of the split and found similar results (Figure S1-4).

To confirm these modeling results, we next investigated the transfer function T(t) that transforms wastewater viral titers to clinical cases. We borrowed this concept from the field of signal processing, where a transfer function represents the mathematical relationship between the numerical input to a dynamic system and the resulting output (Pollock, 2011). While the previous ABC analysis modeled the delay for each individual case to find the distribution of delays, the transfer function analysis focuses on relating the shapes of the wastewater signal and clinical case signal. The shape of T(t) can provide insight on the time delay between the two signals and can reflect factors such as how long it takes someone to request testing and the probability that someone gets a positive test result over the course of their infection (Figure 4A and C).

**Figure 4.**
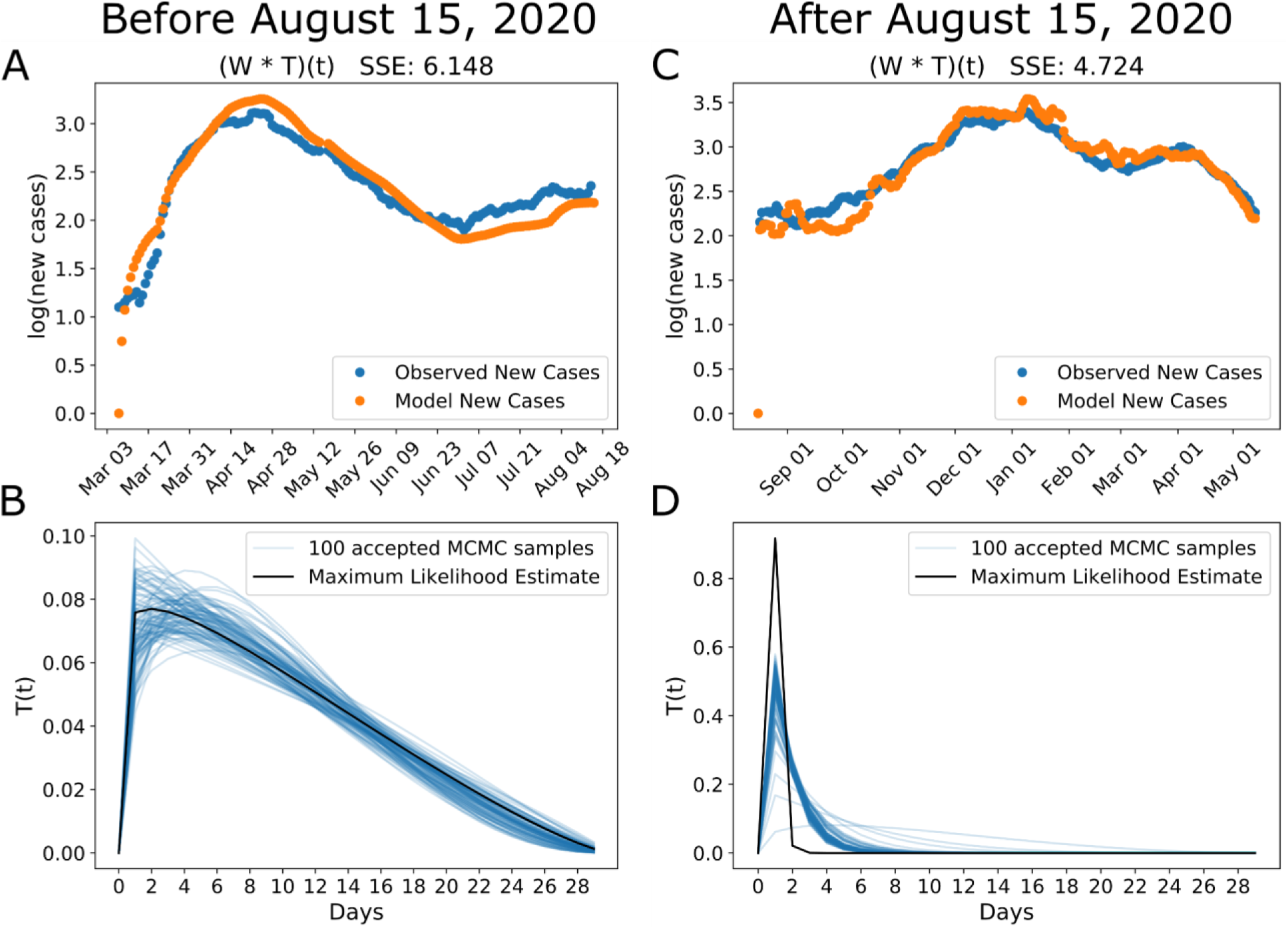
Transfer function between wastewater and clinical cases becomes more peaked in the second wave of the pandemic. We modeled clinically reported new cases as the convolution between wastewater viral titers and an unknown transfer function. (A, C) Our model finds parameters of a beta function that minimizes the sum of squared error (SSE) between the model prediction (orange) and the observed (blue) clinical new cases. (B, D) The maximum likelihood estimate of the transfer function (black) with 100 accepted Markov Chain Monte Carlo (MCMC) parameter sets in blue. Before 8/15, the transfer function has a broad peak and long tail. After 8/15, the transfer function becomes more sharply peaked.

We found that the shape of the transfer function changed between the first and second waves, reflecting changing relationships between infections and clinical testing. Using data before August 15, the inferred transfer function had a broad peak and long tail, with the peak at approximately 3 days and an average of 10 days, which is within the confidence interval of the ABC model (Figure 4B). The broad shape implies that the process of infected individuals getting counted as cases has a broad distribution, with some individuals getting reported very quickly but others taking up to weeks. In this situation, wastewater viral titers could be an early indicator of disease dynamics before clinical test results come back positive. As the pandemic progressed, the inferred transfer function became more sharply peaked around a 1-day time lag, which is consistent with the results of the ABC model. This sharp distribution indicates that wastewater and reported cases track each other closely (Figure 4D). In this case, wastewater viral titers have less utility as an early warning system because increased clinical testing capacity effectively captures new infections in a timely manner.

Taken together, these results suggest that the relationship and time lag between wastewater viral titers and positive clinical tests changed over the course of the pandemic. Wastewater was more effective as a leading indicator in the first wave of the pandemic, and this early warning effect diminished drastically in the second wave, perhaps as clinical testing availability increased. Thus, parameters such as the time lag between wastewater signal and clinical signal and the transfer function describing their relationship could also be used to evaluate clinical test availability or capacity.

### Relationship between wastewater data, new cases, and deaths is different in the first and second waves

Given that wastewater titers correlated with new clinical cases, we next investigated whether we could use wastewater titers to predict COVID-19 deaths. We found that the relationships between wastewater titers, new cases, and deaths were different in the first and second waves, suggesting that wastewater may have a direct, mechanistic relationship with new cases, but an indirect relationship with deaths, indicating that wastewater data must be used in conjunction with other public health datasets for making policy decisions.

In the first surge (Mar-May 2020), both new cases and deaths peaked shortly after wastewater peaked (Figure 5A). In this situation, wastewater titers could be a predictive indicator of COVID-19 deaths. However, in the second surge (Nov 2020-Mar 2021), wastewater viral titers and new cases increased to levels higher than in the first surge, while deaths remained lower than the first wave (Figure 5A). In this situation, wastewater titers were an indicator of new cases but not deaths. This difference could be due to a combination of several factors. First, the first wave strongly affected people in the 80+ year bracket, while the second wave has seen an increase in positivity in the 0-29 year bracket (Wikle et al., 2020). Older adults with comorbidities have higher mortality rates from COVID-19 (Centers for Disease Control and Prevention, n.d.). Second, medical professionals have gained experience in treating and managing the disease since the first wave, thus leading to reduced deaths in the second wave (Q. Liu et al., 2020). Third, changes in human practices, such as improved hygiene and social distancing, could reduce the viral inoculum, resulting in less severe disease (Spinelli et al., 2021). Fourth, the increase in testing means a higher proportion of cases are being diagnosed, possibly including those who are asymptomatic or only mildly ill, so the proportion of deaths to new cases would be lower. This explanation is consistent with the decrease in WC ratio (Figure 2) seen in the second wave. We explored the possibility that variants of concern with increased transmissibility like B.1.1.7 (Frampton et al., 2021; Graham et al., 2021), could have influenced the ratio of deaths to new cases. However, the proportion of B.1.1.7 in the last week of January 2021, during the peak of the second wave, was estimated to make up only an average of ∼2.1% COVID-19 cases in the U.S (Washington et al., 2021). Furthermore, B.1.1.7 did not make up a large proportion of wastewater viral titers in the Boston Area until Feb-March 2021, towards the end of the second wave (Lee et al., 2021).

**Figure 5.**
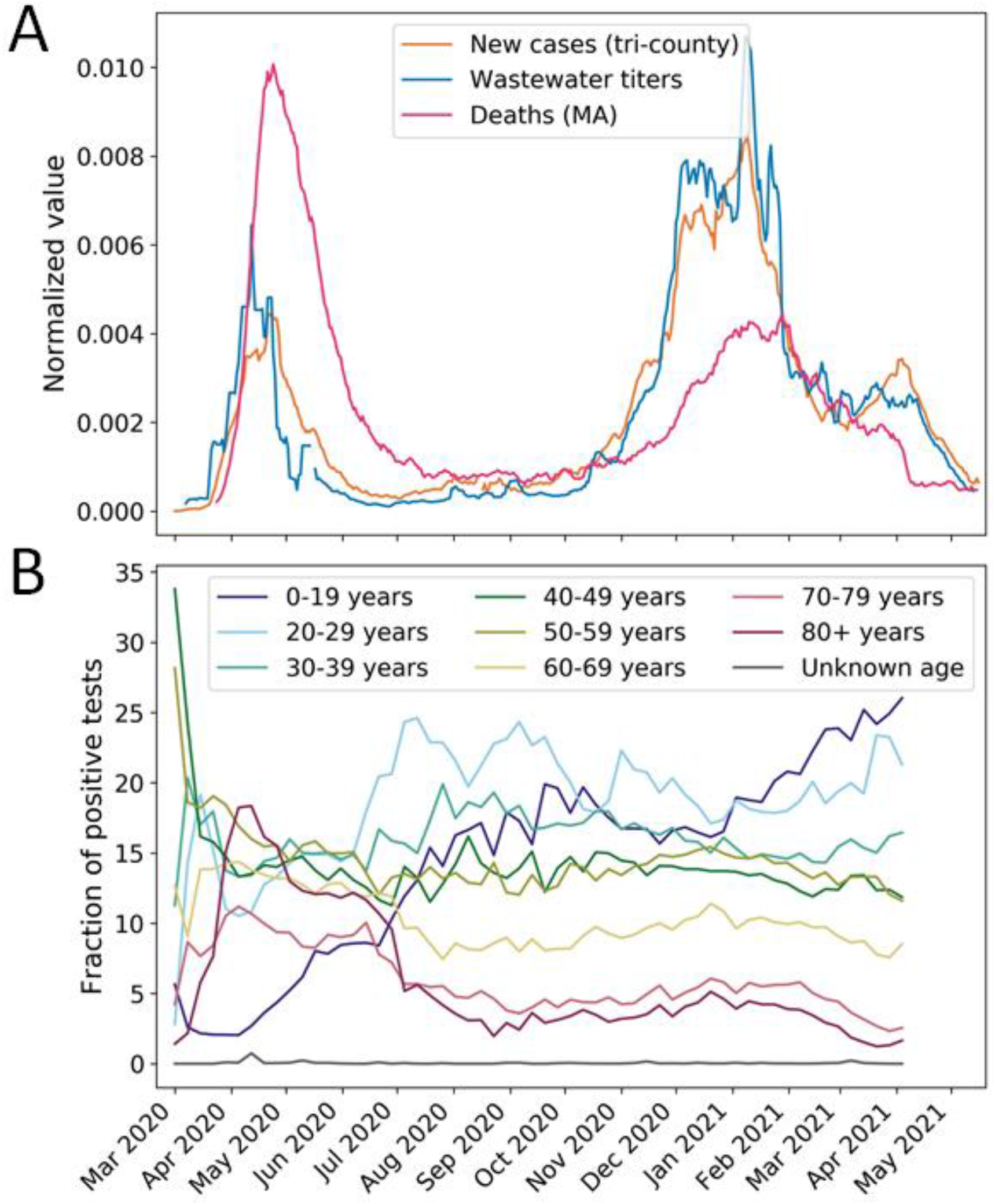
Wastewater is an early warning of new cases but not deaths, perhaps due to a changing demographic of the pandemic. (A) Seven-day averages of wastewater viral titers, new clinical cases in the three counties served by the WWTP (Massachusetts Department of Public Health, 2021b, 2020a, 2020b), and new reported deaths in the state of Massachusetts (Massachusetts Department of Public Health, 2021a). All three datasets are normalized by their sums for comparison. (B) Fraction of positive tests in Massachusetts by age bracket (Massachusetts Department of Public Health, 2021c). Positivity peaked among 80+ year-olds in the first wave of the pandemic, whereas the second wave saw an increase in positivity in 0-29 year-olds.

Notably, wastewater viral titers reflect the number of new cases regardless of the demographics or severity of cases. With the changing demographics of the disease, changes in clinical practice, and increased testing capacity, the power of wastewater to predict new cases and COVID-related deaths also changes. Therefore, wastewater should not be used alone to predict public health outcomes, but rather should be used in combination with other data sources to understand the pandemic and inform decision making.

## Discussion

Many groups have demonstrated that wastewater SARS-CoV-2 titers reflect trends in new COVID-19 cases, and wastewater viral titers have been observed to precede trends from clinical surveillance (Bar-Or et al., 2020; Kocamemi et al., 2020; Medema et al., 2020; Randazzo et al., 2020; Wurtzer et al., 2020). Here, we developed three new metrics to integrate wastewater and clinical data to quantitatively understand their relationship and the public health response. We introduced the ratio of wastewater viral titers to clinical cases (WC ratio) as a simple metric which may reflect testing capacity. We also applied two independent types of modeling to quantify the lead time between wastewater data and clinical data. These models show that wastewater’s lead time changes over the course of the pandemic as public health responses adapt. Finally, we showed that a third public health data stream, COVID-19 deaths, also has changing relationships with cases and wastewater over the pandemic, suggesting that wastewater data cannot be used alone and should be integrated with public health data to make policy decisions. Together, this work demonstrates the utility of combining wastewater surveillance with multiple public health data streams to provide a more nuanced view of the changing public health response to the COVID-19 pandemic.

The ratio between wastewater viral titers and clinical cases (WC ratio) is a useful indicator of the public health response and testing coverage and could be used to gauge the intensity of public health interventions in the face of changing disease incidence. In addition, short-lived trends in increased transmission may be more easily detected when analyzing the WC ratio compared to analyzing wastewater and clinical data independently. For example, the WC ratio showed a small spike in early September and early October. These short-lived spikes could be due to community events, such as university reopening and economic reopening phases. These spikes could also be due to changing demographics of disease. As younger people are more likely to have mild or asymptomatic infections, they are less likely to be captured by clinical testing infrastructure, even when tests are readily available. Therefore, the WC ratio should be considered when interpreting wastewater surveillance data because it may help detect asymptomatic disease transmission among the population.

We also introduced two models that showed that the delay between wastewater data and clinical reporting shrank from an average of 6.2 days to no significant difference between the wastewater and clinical trend after August 15, 2020. In addition, the inferred transfer function between wastewater viral titers and reported cases shifted from a broad to a sharp peak, suggesting quicker access to clinical testing. These results suggest that wastewater surveillance can be useful as an early warning indicator of disease incidence when clinical testing is limited and also as a method to understand ramp up and scale down of community based testing capacity in different phases of the pandemic (Olesen et al., 2021). Importantly, modeling the time lag and transfer function between the wastewater viral titers and clinical cases provides a more quantitative method to understand their relationship and assess the pandemic response.

There could be many factors contributing to the decreasing delay between trends in wastewater and trends in clinical cases. Changing criteria to qualify for clinical testing, individual behavior in requesting tests, availability of convenient testing locations, and lab turnaround time can all affect the time lag between when a patient is infected and when their positive result is reported. In the beginning of the first wave, clinical testing was largely limited to those who met a restrictive combination of symptoms and exposure, gradually expanding to those who had exposure history (Becker, 2020; Brown et al., 2020). However, the initiation of the Massachusetts #StopTheSpread program on July 8, 2020 and its expansion on August 7, 2020 made walkup, on-demand testing available to the public, aiding in the identification of asymptomatic and pre-symptomatic cases (Murphy, 2020; Office of Governor Charlie Baker and Lt. Governor Karyn Polito et al., 2020). Widespread testing by local colleges further expanded this segment of identified cases through fall and winter (Broad Institute, 2020). Reported case counts thus depended heavily on public health resources and policy. Wastewater surveillance is not subject to these social and logistical limitations and can therefore serve as a more instantaneous and unbiased readout of new cases during the pandemic. We and others have shown that wastewater likely detects a short period of high viral shedding early in infection (Hoffmann and Alsing, 2021; Wu et al., 2020a), whereas patients can test positive during PCR testing of respiratory samples for longer periods of time (Wölfel et al., 2020; Zheng et al., 2020), suggesting that wastewater could be more specific to newly infected patients. However, wastewater surveillance does not necessarily provide a readout of hospitalizations or deaths because these numbers also depend on who is infected and their access to healthcare, which cannot be distinguished via wastewater monitoring (Olesen et al., 2021). Therefore, wastewater should be used in conjunction with additional clinical data streams when making public health decisions related to hospitalizations and mortality.

This study has several limitations. First, the interpretation of the WC ratio relies on the assumption that the viral shedding rate did not drastically change over the course of the pandemic. While some variants of SARS-CoV-2 have been reported to have higher shedding rates or longer shedding duration (Frampton et al., 2021; Kissler et al., 2021), the B.1.1.7 variant did not make up a large proportion of wastewater viral titers in the Boston Area until March 2021, well past the periods in the summer where we described the notable peaks in the WC ratio (Lee et al., 2021). There have been mixed reports of the difference in shedding rate between symptomatic and asymptomatic people (Han et al., 2020; Van Vinh Chau et al., 2020; Zhou et al., 2020), but in any case, it is unlikely that the ratio of asymptomatic to symptomatic cases would swiftly change in a catchment of 2.25 million people. Second, there are some fluctuations in the WC ratio throughout summer 2020 and spring 2021 that we were not able to tie back to community events that potentially increased social contacts and disease transmission. These fluctuations could arise from the noise in both datasets and from community behaviors that we did not consider. Private gatherings would be hard to monitor, but perhaps more detailed analysis of mobility data could enhance our interpretations of these fluctuations. Third, in practice, the WC ratio is particularly useful when considering deviations from a stable baseline value. However, assessing such deviations in real time could be difficult, especially in the beginning stages of a pandemic before a baseline is reached. Additionally, it could be difficult to distinguish a shifting baseline from true long-term trends. It is also difficult to use clinical data to verify the silent community spread detected by the WC ratio in the early summer, particularly if those affected were asymptomatic and did not seek hospitalization. In any case, increases in the WC ratio could prompt officials to increase their public health messaging to quell these silent peaks and to remain on alert for further trends.

There are many extensions to this work that will make wastewater surveillance data more integrated and useful for the public health response. For example, wastewater viral titers and clinically reported cases are inherently linked to the number of true infections in the population. More mechanistic modeling is necessary to infer the underlying trend of true infections from the observable wastewater and clinical data (Fernandez-Cassi et al., 2021). Similarly, wastewater viral titers are linked to true infections by a transfer function that describes population-level shedding, while clinical cases are linked to true infections by a transfer function that describes population-level testing availability and turnaround time (Olesen et al., 2021). Inferring these two transfer functions will allow us to understand these parameters separately. Combining wastewater data with clinical and demographic data may also allow us to infer these transfer functions per demographic group, giving us finer resolution understanding of viral shedding parameters and access to testing. As the pandemic has progressed and public health measures have changed, the utility of wastewater surveillance has also changed. In this time-varying context, integrative models can make wastewater data more flexible, useful, and predictive.

## Conclusions

Wastewater is one useful data stream in the public health response toolkit, and its utility can change over time based on the public health response. Here, we introduced a new metric (wastewater viral titer to clinical case ratio, WC ratio) that may be a useful indicator of clinical testing capacity. We also develop methods to describe two ways that the relationship between wastewater viral titers and clinically reported cases changed over the course of the COVID-19 pandemic in Massachusetts. First, there was an approximately 6-day time lag between wastewater signal and clinically reported cases in the first wave, which was not present in the second wave. Second, the transfer function describing the relationship between wastewater and clinical time series became more sharply peaked in the second wave. These results suggest that Massachusetts’s testing capacity increased substantially, with fewer delays and increased testing availability in the second wave and are consistent with state-wide expansion of on-demand clinical testing throughout the summer. Therefore, evaluating the relationships between wastewater and various public health data streams using these new metrics can provide a real-time evaluation of public health responses, and more integrative models can help increase the utility and application of wastewater surveillance for managing the ongoing COVID-19 pandemic as well as future pandemics.

## Methods

### Sample collection and viral inactivation

700 24-hour composite samples of raw sewage were collected from the Deer Island Wastewater Treatment Plant (comprised of northern and southern influents) in Massachusetts from March 4, 2020 to May 13, 2021. The Deer Island Wastewater Treatment Plant serves approximately 2.3 million individuals in the greater Boston area, comprising primarily Norfolk, Suffolk, and Middlesex counties. The average flow rate is 178 million gallons per day (MGD) for the northern influent and 93 MGD for the southern influent. Historical samples from March 4 to March 17, 2020 were stored at 4°C at the WWTP and delivered to the lab. Starting on March 18, the sewage samples were transported to the lab on the same day of sample collection with ice. If the samples were collected on weekends or holidays, they were stored at 4°C and transported to the lab the next business day with ice. Upon receipt, samples were brought to 60°C and pasteurized for 1 hour to inactivate the virus.

### Lab analysis

Samples were analyzed using previously described methods (Wu et al., 2020a). Briefly, samples were filtered to remove large particulate matter using a 0.2uM vacuum-driven filter (EMD-Millipore SCGP00525 or Corning 430320, depending on sample turbidity). We used Amicon Ultra-15 centrifugal ultrafiltration units (Millipore UFC903096) to concentrate 15ml of wastewater approximately 100x. Viral particles in this concentrate were immediately lysed by adding AVL Buffer containing carrier RNA (Qiagen 19073) to the Amicon unit before transfer and >10 minute incubation in a 96-well 2mL block. To adjust binding conditions, 100% ethanol was added to the lysate, and samples were applied to RNeasy Mini columns or RNeasy 96 cassettes (Qiagen 74106 or 74181). RNA samples eluted from the RNeasy kit were subjected to one-step RT-qPCR (ThermoFisher 4444436) analysis in triplicate for N1, N2, and PMMoV amplicons on CFX96 and/or CFX-Connect instruments based on the following protocol: 50°C 10 mins for reverse transcription, 95°C 20 s for RT inactivation and initial denaturation, and 48 cycles of denature (95°C 1 s) and anneal/extend (55°C 30 s). Cts were called from raw fluorescence data using the Cy0 algorithm from the qpcR package (v1.4-1) in R (Guescini et al., 2008), and manually inspected for agreement with the raw traces in the native BioRad Maestro software.

### Data processing

A standard curve was generated using serial dilutions of Twist Bioscience synthetic SARS-CoV-2 RNA control 2 (MN908947.3) and used to convert Ct values into copies per well. We used pepper mild mottle virus (PMMoV) as a fecal indicator and quantified it relative to SARS-CoV-2 in each sample using the standard curve for N1, as synthetic RNA for PMMoV was not available and our PMMoV normalization method is dependent on ratios of PMMoV values rather than absolute values. Ct values above 40 were considered as non-detect. The copies per well were multiplied by a dilution factor accounting for the volume changes described above (RNA extraction, concentration, etc.) and then divided by the original sewage volume (15 ml) to convert to a sewage concentration (copies per liter). Concentrations of N1 and N2 replicates were averaged first within each primer set and then across primers to get the final SARS-CoV-2 concentration; replicates of the PMMoV amplicon were averaged. Samples were required to have at least two quantified replicates between N1 or N2, and at least one detected PMMoV replicate to be considered a detection. For median normalization using PMMoV, SARS-CoV-2 concentrations were divided by the PMMoV levels and multiplied by a reference PMMoV value derived as the median of our dataset. Data on the concentration of SARS-CoV-2 viral RNA in Massachusetts wastewater is publicly available at https://www.mwra.com/biobot/biobotdata.htm.

### Clinical case analysis

We downloaded clinical case data from March 1, 2020 to May 16, 2021 from Norfolk, Suffolk, and Middlesex Counties from Mass.gov (Massachusetts Department of Public Health, 2021b, 2020a, 2020b). Because Mass.gov county-level reporting in August 2020 did not include data before mid-April 2020, we combined the first two files starting on April 18, 2020. We summed the clinical cases from each county to represent the cases in the catchment of the wastewater treatment plant and calculated the new cases per day. We downloaded positive test rates by age and state level death data from Mass.gov (Massachusetts Department of Public Health, 2021b, 2021a).

### Approximate Bayesian computation for estimating delay distribution

We used approximate Bayesian computation (ABC) to find the delay distribution between when an infected individual’s viral shedding appears in wastewater and when they are counted as a clinical case before and after August 15, 2020. We first normalized wastewater viral titers by their sum for each portion of the data (before/after August 15, 2020). We assumed the delay δ is normally distributed with mean μ and standard deviation σ. We chose prior distributions for μ and σ where μ ∼ Norm(0,10) and σ ∼ Exp(0.1). We sampled 10,000 values of μ and σ from the priors. For each iteration, we sampled a delay for each clinically reported case. We generated simulated wastewater data by adding the delay for each case to the date they were actually reported. We summed the number of delayed cases per day and normalized by the sum. We calculated the average per day sum of squared errors (SSE) between the simulated wastewater data and the actual wastewater data. We accepted the values of μ and σ if the average SSE was less than some cutoff value ε. We tuned ε so approximately 10-20% of the iterations were accepted.

### Convolution to estimate the transfer function

We used a similar approach as we previously reported to find the transfer function that describes the relationship between the shape of the wastewater data and clinical data (Wu et al., 2020a). Because we were interested in the shapes rather than the magnitudes of the curves, we divided the wastewater data by the median ratio between wastewater and clinical data to get both curves on the same scale. We model the clinically reported cases C(t) as the convolution between the scaled wastewater data W(t) and the unknown transfer function T(t) before and after August 15, 2020: log10(C(t)) = log10([T * W](t)). We hypothesized that the transfer function could be fit by a beta distribution with parameters α, β, and scaling factor c because beta distributions harbor a rich variety of shapes with only two parameters. We defined the score function as the sum of squared errors (SSE) between log10(clinically reported cases) and log10([T * W](t)). We used the L-BFGS method in the scipy.optimize.minimize function to find parameters α, β, c of the beta distribution that minimized the SSE. For initial parameter guesses, we used a combination of α = [2, 20, 50, 100, 200], β = [2, 20, 50, 100, 200], and c = [0.01, 0.1], which gives a wide variety of starting shapes for the transfer function.

### Markov Chain Monte Carlo (MCMC) simulation to quantify uncertainty in transfer function

Under the assumption of normally distributed errors, minimizing the SSE is equivalent to maximizing the likelihood. We next used MCMC simulation to investigate the uncertainty landscape around the maximum likelihood estimation of the parameters for the inferred transfer function. Briefly, we started at the maximum likelihood estimate for each parameter α, β, and c. We defined the transition function as a normal distribution centered around the previous parameters, with standard deviation (1, 1, 0.001) for α, β, and c, respectively. At each iteration, we selected a new set of parameters using the transition function and computed the log likelihood. We always accepted the new parameters if the log likelihood was higher. If the log likelihood was lower for the new parameters, we accepted them with probability exp(-delta(SSE)). We selected 100 random accepted parameter sets and plotted them in Figure 4B and 4D to illustrate the uncertainty around the maximum likelihood estimate of the transfer function. MCMC simulation was done with python 3.6.5, numpy 1.14.3, pandas 0.23.0, and scipy 1.1.0.

## Data Availability

Data on the concentration of SARS-CoV-2 viral RNA in Massachusetts wastewater is publicly available at https://www.mwra.com/biobot/biobotdata.htm.

## Acknowledgments

We thank the Deer Island wastewater treatment facility team for providing the samples for analysis, Penny Chisholm (MIT) and Allison Coe (MIT) for access to equipment and other supplies, Mathilde Poyet (MIT) and Shandrina Burns (MIT) for logistical support, and Shijie Zhao (MIT), Ethan Evans (MIT), Yanjia Jason Zhang (MIT), Scott Olesen, Stephen Estes-Smargiassi (MWRA), and Stefan Wuertz (NTU) for helpful discussion. We express our deep gratitude to all healthcare professionals and first-line responders who have been caring for patients with COVID-19.

This work was supported by the Center for Microbiome Informatics and Therapeutics and Intra-CREATE Thematic Grant (Cities) grant NRF2019-THE001-0003a to JT and EJA; National Institute on Drug Abuse of the National Institutes of Health award numbers K23DA044874 to PRC; and R44DA051106 to MM and PRC, Hans and Mavis Psychosocial Foundation funding, and e-ink corporation funding to PRC; funds from the Massachusetts Consortium on Pathogen Readiness and China Evergrande Group to TBE, PRC, MM, and EJA. The content is solely the responsibility of the authors and does not necessarily represent the official views of the funding institutions.

