## Supplemental for "Metrics to relate COVID-19 wastewater data to clinical testing dynamics"

**Supplemental Table S1: Four Phases of Massachusetts Reopening**

**Phase 1 (effective May 18, 2020)**

In the first phase, manufacturing and construction can reopen provided they follow standards meant to curb the spread of the virus. Houses of worship can resume services if they follow social distancing. Outdoor services are encouraged. Hospitals and community health centers are allowed to provide high-priority preventative care, pediatric care and treatment for high risk patients and conditions. (Baker, 2020a)

**Phase 2 (Step 1 effective June 8, 2020; Step 2 effective June 22, 2020)**

*Effective at least three weeks after Phase 1:* Retail businesses, restaurants, hotels and other personal services such as nail salons and day spas can reopen with restrictions. Hospitals and community health centers are allowed to provide less-urgent preventative care, including teeth cleanings and certain elective procedures. More recreation is allowed to restart, including campgrounds, playgrounds, public pools, athletic fields and courts and youth sports in a limited fashion. (Baker, 2020a, 2020b, 2020c)

**Phase 3 (Step 1 effective July 6, 2020; Step 2 effective October 5, 2020; rollback to Step 1 on December 13, 2020; Step 2 effective again March 1, 2021)**

*Effective at least three weeks after Phase 2:* Bars, casinos, gyms, museums and others in the entertainment and arts industries can reopen. All other business activities can resume except for nightclubs and large venues. More recreation is allowed to restart, including youth sorts with games and tournaments, though crowd sizes will be limited. (Baker, 2021a, 2020e, 2020d, 2020f)

**Phase 4 (Step 1 effective March 22, 2021; Step 2 effective May 10, 2021)**

*Effective upon the development of vaccines and treatment:* Full resumption of activity in the "new normal," including travel, all outdoor recreation and activities as well as events in large venues and nightclubs. (Baker, 2021b; Governor's Press Office et al., 2021)

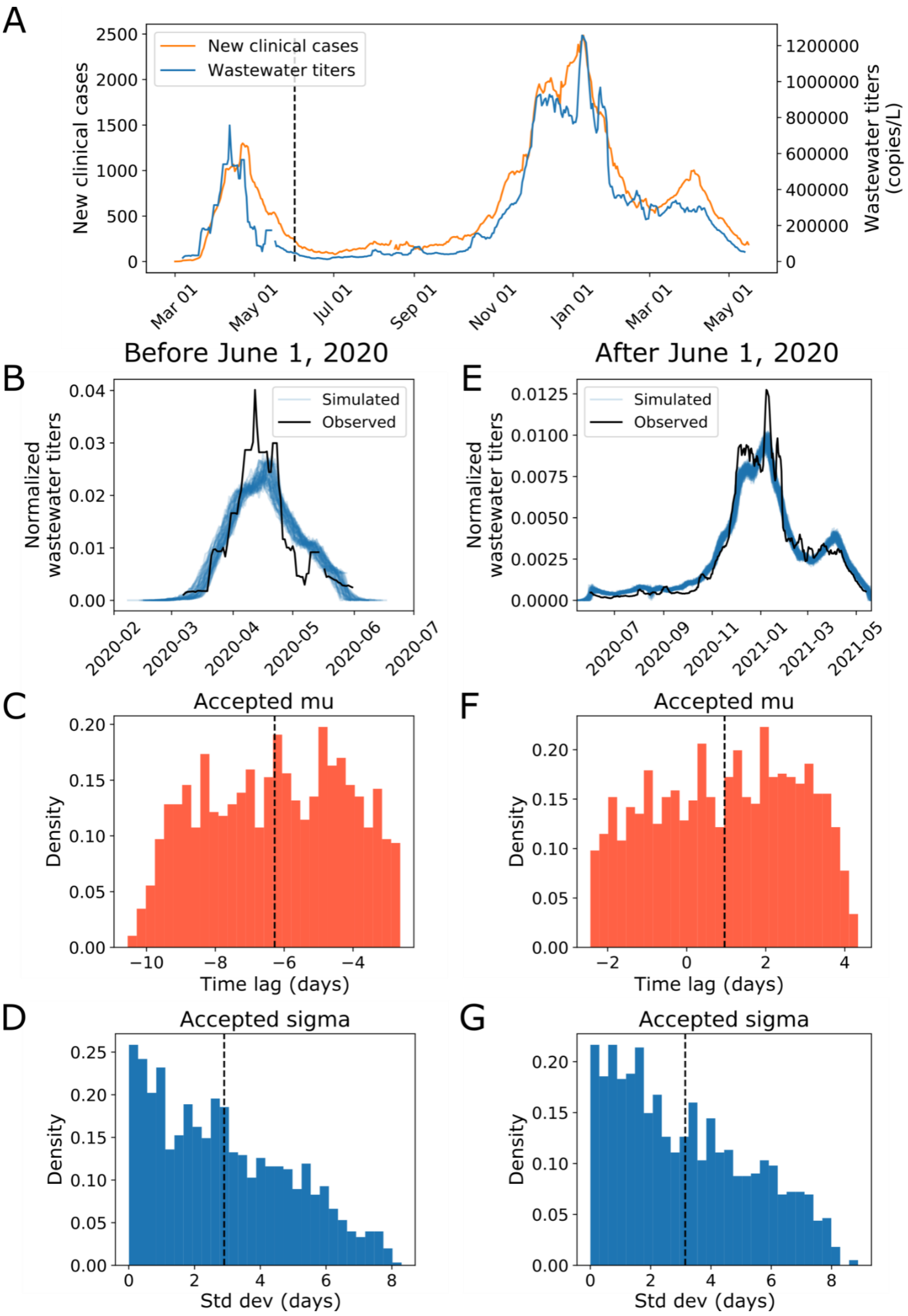

A
New clinical cases
Wastewater titers
New clinical cases
Wastewater titers (copies/L)
Mar 01
May 01
Jul 01
Sep 01
Nov 01
Jan 01
Mar 01
May 01
Before June 1, 2020
After June 1, 2020
B
E
Simulated
Observed
Normalized wastewater titers
C
F
Accepted mu
Density
Time lag (days)
D
G
Accepted sigma
Density
Std dev (days)

**Figure S1 Modeling reveals that time delay between clinical case reporting and wastewater data changes over the course of the pandemic.** We use Approximate Bayesian Computation to determine the distribution of the time lag between when a case shows up in the wastewater surveillance data and when they are clinically reported. (A) Seven-day averages of wastewater data and new clinical case data from Mass.gov shown on a linear scale (Massachusetts Department of Public Health, 2021b, 2020a, 2020b). (B, C, D) Modeling results on data before June 1, 2020. (B) Simulated vs observed wastewater viral titers for data before 6/1. (C) Accepted values for the mean time lag (-6.3 days, 95% CI: -9.8, -2.9) and (D) standard deviation (2.9 days, 95% CI: 0.1, 7.3) of the time lag for data before 6/1. We used 10,000 iterations with a distance threshold of 2.5e-5 and 10.9% of parameter sets were accepted. (E, F, G) Modeling results on data from June 1, 2020 and after. (E) Simulated vs observed wastewater viral titers for data after 6/1. (F) Accepted values for the mean time lag (0.96 days, 95% CI: -2.2, 3.9) and (G) standard deviation (3.2 days, 95% CI: 0.1, 7.6) of the time lag for data after 6/1. We used 10,000 iterations with a distance threshold of 7.5e-7 and 13.1% of parameter sets were accepted. Negative time lags indicate that wastewater signal precedes clinical case reporting and vice versa.

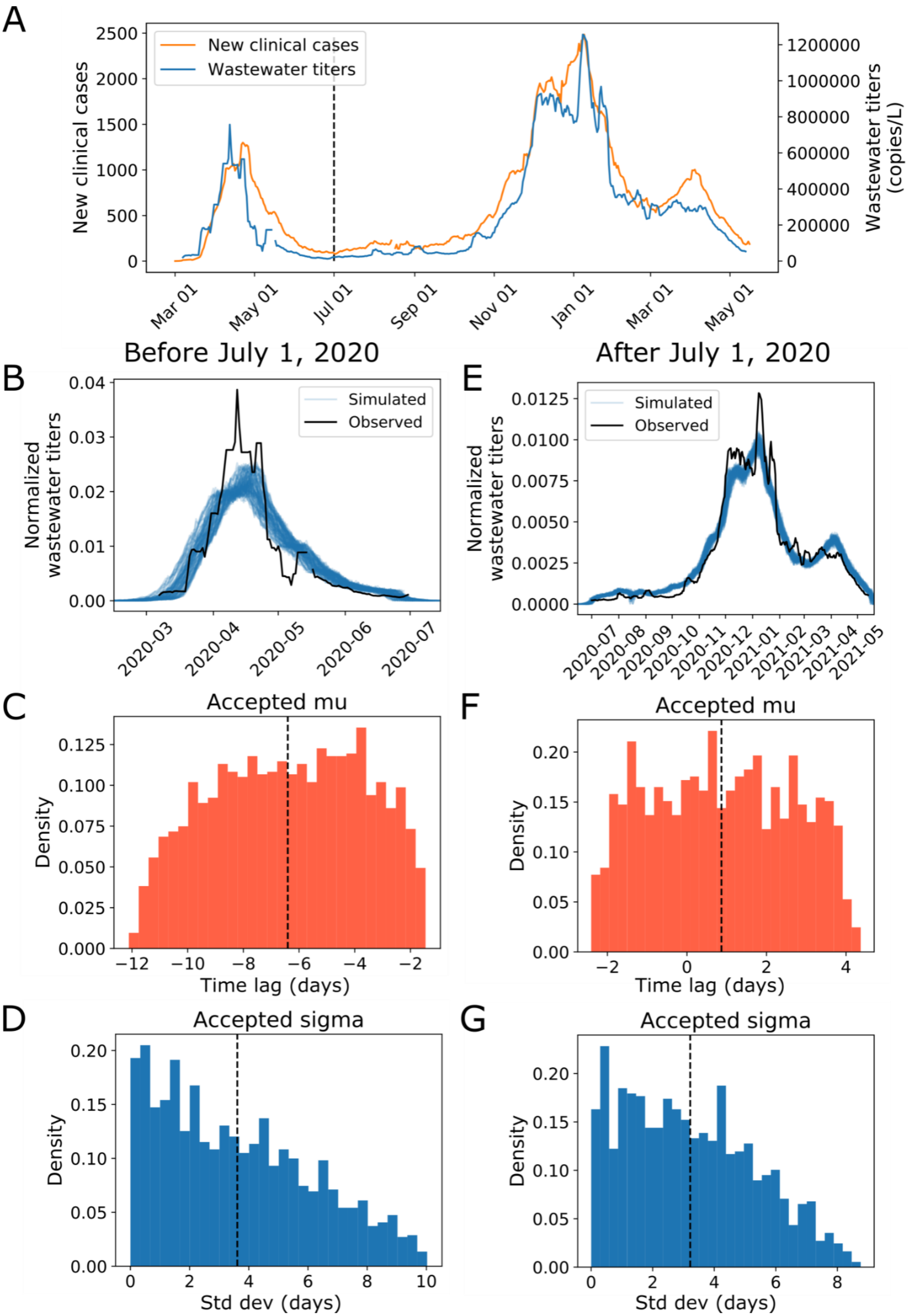

A
New clinical cases
Wastewater titers
New clinical cases
Wastewater titers (copies/L)
Mar 01
May 01
Jul 01
Sep 01
Nov 01
Jan 01
Mar 01
May 01
Before July 1, 2020
After July 1, 2020
B
E
Simulated
Observed
Normalized wastewater titers
C
F
Accepted mu
Density
Time lag (days)
D
G
Accepted sigma
Density
Std dev (days)

**Figure S2 Modeling reveals that time delay between clinical case reporting and wastewater data changes over the course of the pandemic.** We use Approximate Bayesian Computation to determine the distribution of the time lag between when a case shows up in the wastewater surveillance data and when they are clinically reported. (A) Seven-day averages of wastewater data and new clinical case data from Mass.gov shown on a linear scale (Massachusetts Department of Public Health, 2021b, 2020a, 2020b). (B, C, D) Modeling results on data before July 1, 2020. (B) Simulated vs observed wastewater viral titers for data before 7/1. (C) Accepted values for the mean time lag (-6.4 days, 95% CI: -11.2, -2.9) and (D) standard deviation (3.6 days, 95% CI: 0.2, 9.0) of the time lag for data before 7/1. We used 10,000 iterations with a distance threshold of 2.2e-5 and 17.7% of parameter sets were accepted. (E, F, G) Modeling results on data from July 1, 2020 and after. (E) Simulated vs observed wastewater viral titers for data after 7/1. (F) Accepted values for the mean time lag (0.87 days, 95% CI: -2.1, 3.9) and (G) standard deviation (3.2 days, 95% CI: 0.2, 7.5) of the time lag for data after 7/1. We used 10,000 iterations with a distance threshold of 8e-7 and 12.6% of parameter sets were accepted. Negative time lags indicate that wastewater signal precedes clinical case reporting and vice versa.

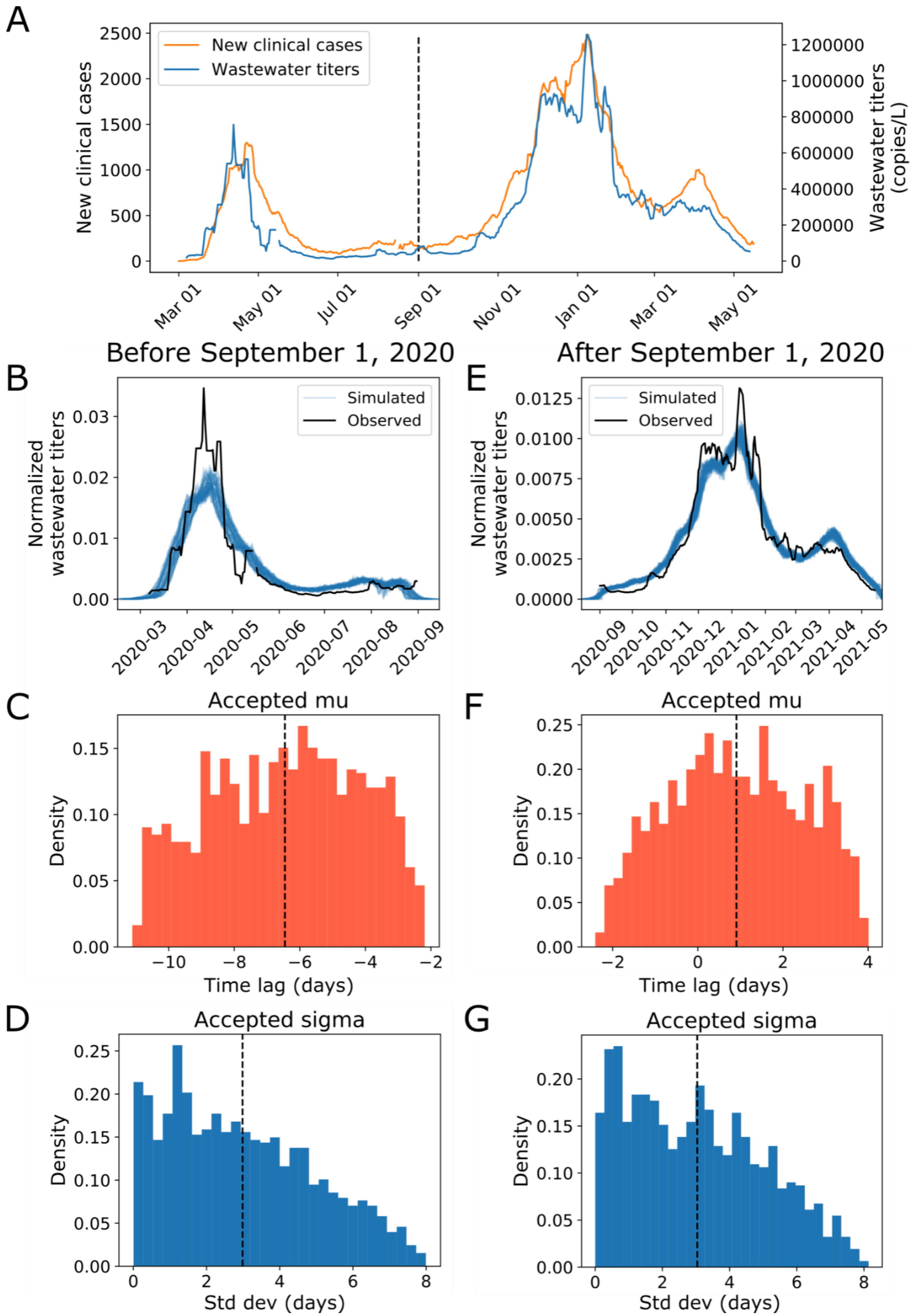

A
New clinical cases
Wastewater titers
New clinical cases
Wastewater titers (copies/L)
Mar 01
May 01
Jul 01
Sep 01
Nov 01
Jan 01
Mar 01
May 01
Before September 1, 2020
After September 1, 2020
B
E
Simulated
Observed
Normalized wastewater titers
C
F
Accepted mu
Density
Time lag (days)
D
G
Accepted sigma
Density
Std dev (days)

**Figure S3 Modeling reveals that time delay between clinical case reporting and wastewater data changes over the course of the pandemic.** We use Approximate Bayesian Computation to determine the distribution of the time lag between when a case shows up in the wastewater surveillance data and when they are clinically reported. (A) Seven-day averages of wastewater data and new clinical case data from Mass.gov shown on a linear scale (Massachusetts Department of Public Health, 2021b, 2020a, 2020b). (B, C, D) Modeling results on data before September 1, 2020. (B) Simulated vs observed wastewater viral titers for data before 9/1. (C) Accepted values for the mean time lag (-6.4 days, 95% CI: -10.5, -2.7) and (D) standard deviation (3.0 days, 95% CI: 0.1, 7.2) of the time lag for data before 9/1. We used 10,000 iterations with a distance threshold of 1.15e-5 and 12.3% of parameter sets were accepted. (E, F, G) Modeling results on data from September 1, 2020 and after. (E) Simulated vs observed wastewater viral titers for data after 9/1. (F) Accepted values for the mean time lag (0.91 days, 95% CI: -1.9, 3.6) and (G) standard deviation (3.0 days, 95% CI: 0.1, 7.2) of the time lag for data after 9/1. We used 10,000 iterations with a distance threshold of 9.5e-7 and 11.5% of parameter sets were accepted. Negative time lags indicate that wastewater signal precedes clinical case reporting and vice versa.

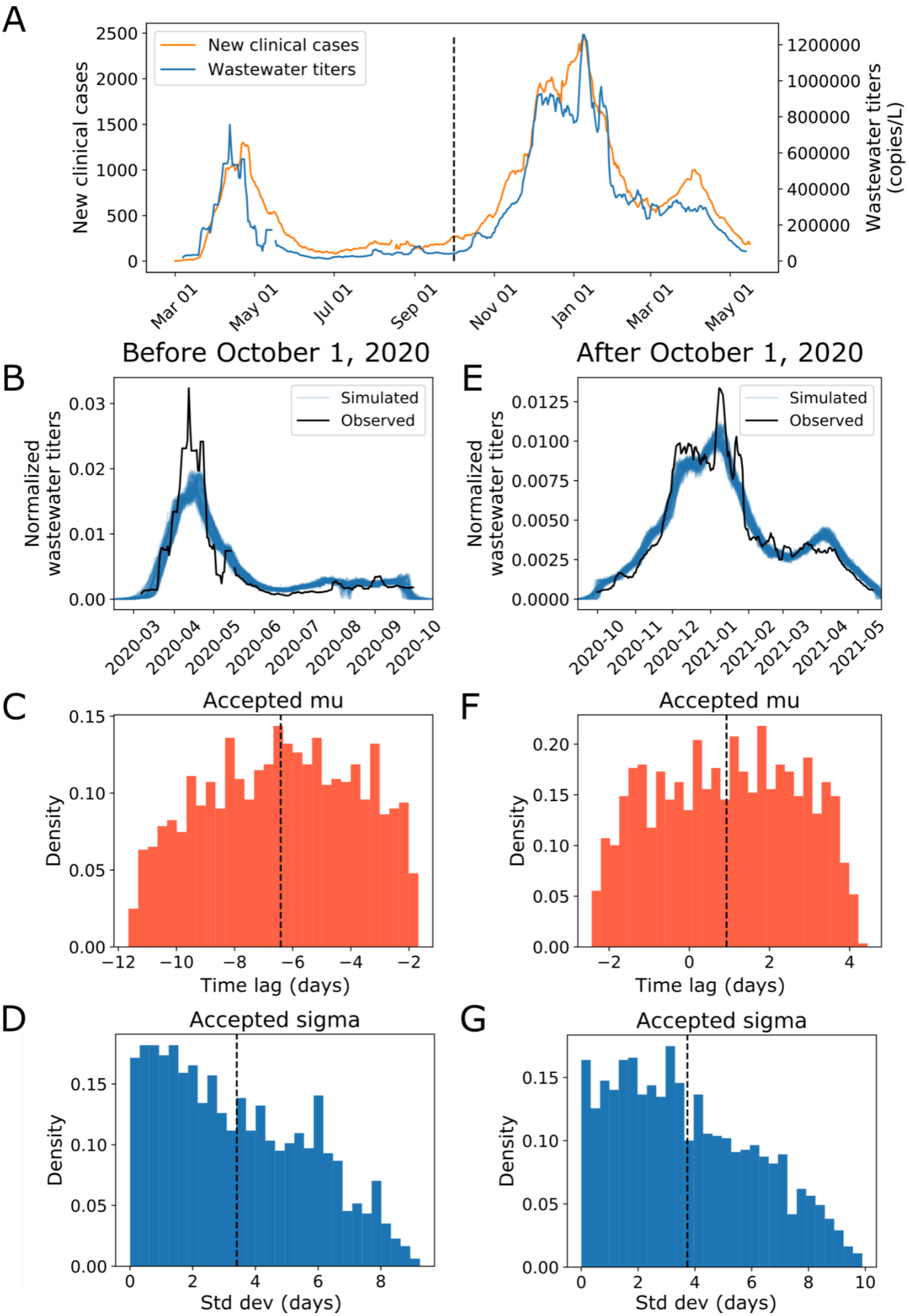

A
New clinical cases
Wastewater titers
New clinical cases
Wastewater titers (copies/L)
Mar 01
May 01
Jul 01
Sep 01
Nov 01
Jan 01
Mar 01
May 01
Before October 1, 2020
After October 1, 2020
B
E
Simulated
Observed
Normalized wastewater titers
C
F
Accepted mu
Density
Time lag (days)
D
G
Accepted sigma
Density
Std dev (days)

**Figure S4 Modeling reveals that time delay between clinical case reporting and wastewater data changes over the course of the pandemic.** We use Approximate Bayesian Computation to determine the distribution of the time lag between when a case shows up in the wastewater surveillance data and when they are clinically reported. (A) Seven-day averages of wastewater data and new clinical case data from Mass.gov shown on a linear scale(Massachusetts Department of Public Health, 2021b, 2020a, 2020b). (B, C, D) Modeling results on data before October 1, 2020. (B) Simulated vs observed wastewater viral titers for data before 10/1. (C) Accepted values for the mean time lag (-6.4 days, 95% CI: -11.0, -2.1) and (D) standard deviation (3.4 days, 95% CI: 0.1, 8.0) of the time lag for data before 10/1. We used 10,000 iterations with a distance threshold of 9.5e-6 and 15.7% of parameter sets were accepted. (E, F, G) Modeling results on data from October 1, 2020 and after. (E) Simulated vs observed wastewater viral titers for data after 10/1. (F) Accepted values for the mean time lag (0.93 days, 95% CI: -2.1, 3.9) and (G) standard deviation (3.5 days, 95% CI: 0.2, 8.0) of the time lag for data after 10/1. We used 10,000 iterations with a distance threshold of 1.1e-6 and 12.6% of parameter sets were accepted. Negative time lags indicate that wastewater signal precedes clinical case reporting and vice versa.
